# Deep learning with multiscale spatial context improves global dengue suitability mapping

**DOI:** 10.64898/2026.08.10.26359943

**Authors:** Carlin Foka Takamgno, Jenicca Poongavanan, Moritz U.G. Kraemer, Tulio de Oliveira, Elizaveta Semenova, Houriiyah Tegally

## Abstract

Infectious-disease risk models often rely on occurrence records that are incomplete and spatially biased by surveillance effort, diagnostic access, and outbreak history. Ecological niche modelling (ENM) can identify areas where disease occurrence is environmentally plausible, yet most approaches represent locations using only pointwise covariate values and therefore overlook the surrounding spatial context and rely on presence-only data. Here, we present a deep-learning framework for presence-only data that estimates relative disease suitability by comparing the environmental conditions surrounding reported occurrences with those sampled across the wider study area. The model processes gridded environmental patches at local, neighbourhood, and broader landscape scales, learns the contribution of each scale, and accommodates missing raster values. Using dengue virus as a global case study, we evaluate whether multiscale spatial representation improves upon point-based ENM baselines including random forest and maximum entropy (MaxEnt) under a spatially disjoint train-test design. The model achieved a Boyce index of 0.971 and an AUC of 0.976 on the held-out test set. Learned scale weights and ablation experiments indicated that neighbourhood context contributed most strongly, while local and broader-scale information provided complementary predictive signals. Compared with point-based baselines, the model identified 6–18% more environmentally suitable area across South Asia, Southeast Asia, and South America, encompassing tens of millions of residents. These findings demonstrate that multiscale spatial context can improve estimates of relative dengue suitability. More broadly, mask-aware convolutional density-ratio estimation provides a flexible framework for mapping environmentally structured pathogens from incomplete, presence-only occurrence data.

## 1 Introduction

Climate-sensitive infectious diseases remain a major global-health challenge because the conditions that support transmission are changing unevenly across space^1–3^. Dengue is a clear example: its geography is increasingly shaped by changing environments, expanding human settlements, and the redistribution and expansion of *Aedes* mosquito vectors, yet its observed distribution remains an imperfect proxy for where transmission is plausible or already occurring^4–8^. Reported dengue occurrences are shaped not only by ecological conditions, but also by surveillance intensity, diagnostic access, reporting practices, and outbreak history^8–10^. A central task in dengue geography and more broadly in the mapping of climate-sensitive infections is therefore not simply to interpolate known cases, but to estimate the latent suitability of locations for disease occurrence: the relative support that ecological, demographic, and vector-related conditions provide for the pathogen-vector-host system^2,7,11^.

Ecological niche modelling (ENM) also known as species distribution modelling (SDM) offers a natural framework for this problem. Rather than treating observed cases as a complete map of transmission, ENM estimates environmental suitability by learning associations between occurrence patterns and the climatic, ecological, demographic and socioeconomic conditions under which they arise^2,11,12^. In infectious-disease applications, this framework has been used to map the potential distribution of pathogens, vectors and reservoir hosts, including for dengue, malaria and West Nile virus, particularly where surveillance is incomplete or spatially biased^4,11,13,14^. Yet, ENM-based disease distribution modelling remains dominated by established presence-only and presence-background frameworks, implemented through algorithms such as Maximum Entropy (MaxEnt), boosted regression trees, generalized additive models and random forests (RF)^15–19^.

A key limitation of these established ENM-based disease distribution models is that they usually represent each occurrence or background location as a vector of covariate values extracted at that point. This pointwise representation is convenient, but it ignores spatial environmental structure around each location^12,20^. For dengue and other pathogens, this can be restrictive. Suitability is unlikely to depend only on the temperature, precipitation, vegetation or urban value at a single pixel. It may also depend on neighbourhood-scale configurations of climate, land surface, water availability, vegetation, human settlement and vector habitat^6,7,16,21^. These considerations motivate a spatially explicit modelling hypothesis: dengue suitability may be better represented as a multiscale environmental context than as a set of focal-pixel covariates.

In biodiversity modelling, deep learning approaches such as convolutional neural networks (CNNs) have recently been proposed as a way to move beyond pointwise covariate extraction by learning directly from spatial environmental neighbourhoods. CNN-based species distribution models can use gridded covariate patches around each occurrence location, allowing the model to learn spatial structure rather than relying only on focal-pixel values or manually specified landscape summaries^20,22,23^. However, this form of spatial representation learning remains largely undeveloped in ENM-based infectious-disease distribution modelling, where recent reviews indicate continued reliance on conventional ENM and machine-learning approaches^15,16^.

Here, we introduce a multiscale convolutional density-ratio ecological niche modelling framework (multiscale CNN-DRE) for infectious-disease distribution mapping, using dengue virus as a test case. This model estimates relative environmental suitability from presence-background data while learning from the gridded conditions surrounding each location, rather than from covariate values at the focal point alone. It considers environmental context at fine, neighbourhood and broader spatial scales and learns how much each scale contributes to suitability at each location, allowing the relevant spatial scale to vary across the study area. The model also accounts explicitly for missing raster values, enabling incomplete covariate data to be used without discarding observations. We compare the proposed framework with point-based baselines, including MaxEnt, random forests and a multilayer perceptron (MLP-DRE), a standard feedforward neural network that implements classifier-based density-ratio estimation using covariate values extracted at individual locations rather than from spatial patches.

We apply this framework to ask whether spatial representation learning improves ENM-based dengue suitability modelling, whether dengue suitability is better captured as a multiscale signal than as a pointwise covariate response, and which spatial scales and covariates drive model predictions. In doing so, we provide both an updated global analysis of dengue suitability and a modelling approach for CNN-based infectious-disease ENM from biased presence-background data.

## 2 Results

In this work, we investigated how spatial context and spatial scale structure the current global suitability of dengue virus. To do this, we developed a deep-learning density-ratio framework that estimates suitability from masked environmental raster patches. The model combines mask-aware partial-convolution feature extraction with a classifier-based density-ratio estimation head, allowing suitability to be learned by contrasting observed presence locations with background locations representing available environmental conditions, while preserving spatial information across incomplete patches. We first compared alternative DRE formulations and identified the best-performing approach, then examined fixed spatial extents to determine which single scale was most informative, and finally built an adaptive multi-scale CNN-DRE to integrate local, neighbourhood, and broader context. We benchmarked this framework against several baselines including MaxEnt, Random Forest and a multilayer perceptron density-ratio estimator (MLP-DRE) using spatial cross-validation and an independent held-out test set.

At the global scale, the multiscale CNN-DRE model outperformed the point-based baseline models on the spatially disjoint held-out test set (Fig. 1G). It achieved an AUC of 0.976 and a Boyce index of 0.971, compared with 0.944 and 0.898, respectively, for the Random Forest baseline and 0.926 and 0.928 for the MLP-DRE baseline. Internal cross-validation supported the discrimination advantage of the multiscale CNN-DRE, which showed the highest median AUC across folds. Cross-validated Boyce index values were more variable and overlapped among models, with the MLP-DRE showing the highest median. The multiscale CNN-DRE model also outperformed MaxEnt on the held-out test set (Boyce = 0.906, AUC = 0.931; Supplementary Fig. S1).

**Figure 1.**
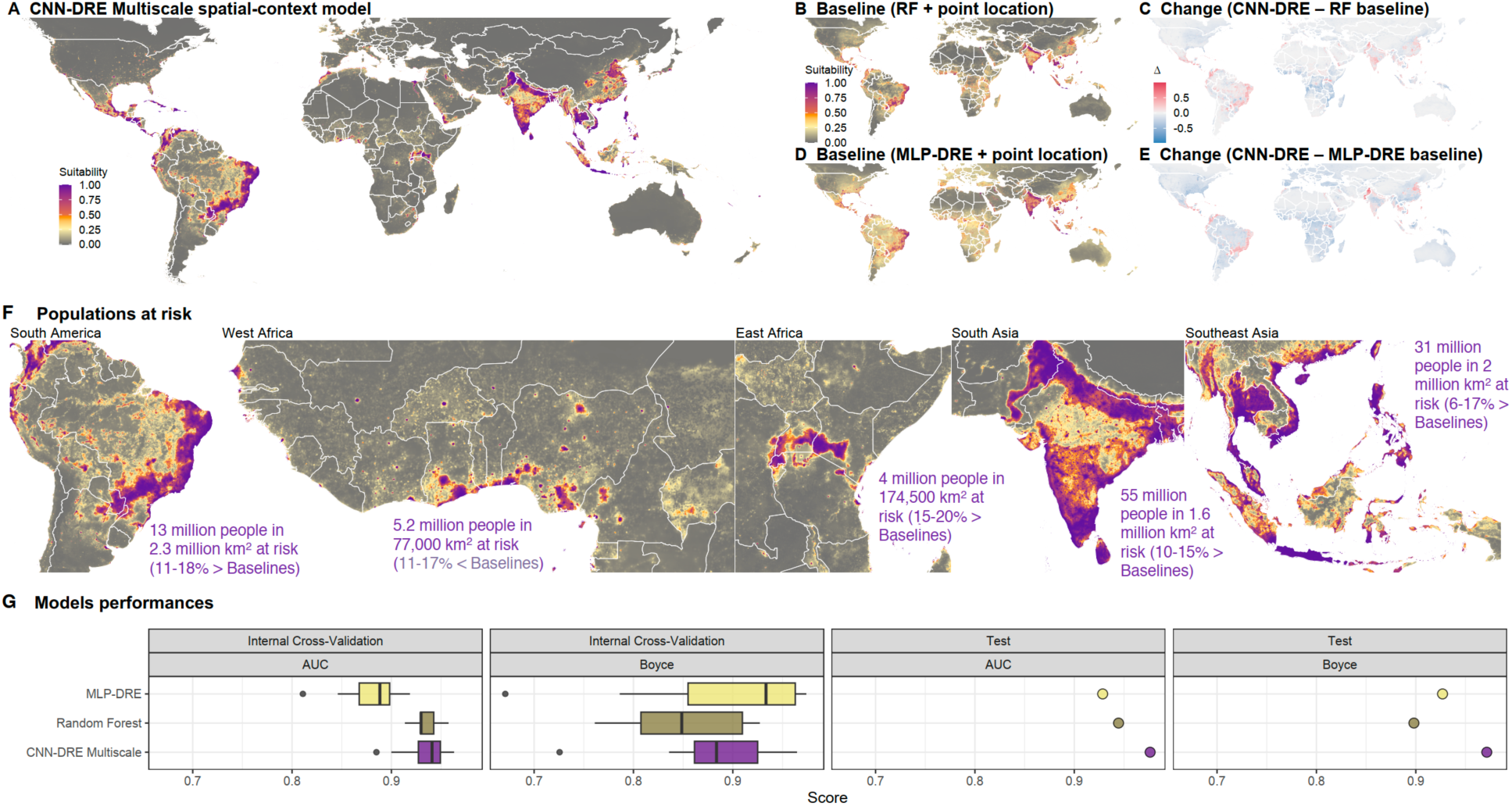
Multiscale spatial context increases predicted suitability and estimated populations at risk. (A) Global suitability predicted by the multiscale CNN-DRE model. (B, D) Baseline suitability predictions from a Random Forest model and an MLP-DRE model using point-location information. (C, E) Difference maps showing the change in suitability between the multiscale CNN-DRE model and each baseline. Positive values indicate higher suitability under the multiscale CNN-DRE. (F) Regional zooms showing populations and land area classified as at risk in South America, West Africa, East Africa, South Asia, and Southeast Asia. The multiscale CNN-DRE identifies additional at-risk populations and areas relative to the baselines, with the largest exposed population in South Asia. (G) Internal cross-validation and held-out test performance, measured by AUC and Boyce score, show that the CNN-DRE multiscale model performs better than or comparably to the Random Forest and MLP-DRE baselines.

The performance gain was accompanied by geographically structured differences in predicted dengue suitability. Relative to the Random Forest and MLP-DRE point-location baselines, the multiscale CNN-DRE preserved the main endemic cores while redistributing suitability across regional margins and fragmented transition zones, particularly in South America, West Africa, East Africa, South Asia and Southeast Asia (Fig. 1A-E). These differences altered the surface area classified as at risk (Fig. 1F). In South America, the multiscale CNN-DRE classified approximately 2.3 million km² as at risk, compared with 1.95–2.07 million km² across the two point-based baselines, representing an increase of 11–18%. The corresponding estimates were 174,500 km² versus 145,000–152,000 km² in East Africa (15–20% higher), 1.6 million km² versus 1.39–1.45 million km² in South Asia (10–15% higher), and 2 million km² versus 1.71–1.89 million km² in Southeast Asia (6–17% higher). In contrast, our multiscale CNN-DRE classified 77,000 km² as at risk in West Africa, compared with approximately 86,500– 92,800 km² for the baselines, representing an 11–17% reduction. The areas identified by the multiscale CNN-DRE contained an estimated 13 million people in South America, 5.2 million in West Africa, 4 million in East Africa, 55 million in South Asia and 31 million in Southeast Asia.

To assess whether these performance differences described above were robust to background selection, we repeated the model comparison using an alternative sampling strategy. The main analysis used target-group background points to account for spatial variation in sampling effort, whereas the sensitivity analysis used uniformly sampled background points while retaining the same spatial cross-validation design and procedure for constructing the disjoint held-out test set. Under uniform-background sampling, the multiscale CNN-DRE achieved an AUC of 0.948 and a Boyce index of 0.938 (Supplementary Fig. S1A). Its AUC was comparable to that of Random Forest (0.947) and higher than those of the MLP-DRE (0.910) and MaxEnt (0.901), while its Boyce index exceeded all three baselines (0.781, 0.839 and 0.907, respectively). Thus, the relative performance of the multiscale CNN-DRE was not specific to the use of target-group background points.

Beyond testing sensitivity to background selection, we next examined whether the advantage of the multiscale CNN-DRE arose from its use of spatial arrangement rather than simply from the additional covariate values contained within patches. For this purpose, we trained and evaluated an otherwise identical shuffled-patch control using the same spatial folds and held-out test set. Spatial positions were permuted within each scale-specific patch while preserving the covariate content and associated validity masks. Shuffling reduced the AUC from 0.976 to 0.956 and the Boyce index from 0.971 to 0.876 (Supplementary Fig. S1B). Although the shuffled model retained substantial discriminatory ability, the decline in both metrics indicates that the unshuffled multiscale CNN-DRE used information contained in the spatial configuration of covariates.

To better understand the spatial basis of these predictions, we examined how local, neighbourhood and broader spatial context contributed to the multiscale CNN-DRE. The global dominant-scale map showed clear geographic variation in scale use, indicating that the model did not apply a single uniform spatial extent across the study area (Fig. 2A). However, scale use was strongly skewed towards local and neighbourhood context rather than broader regional context. Across all land pixels, the location-specific fusion weights generated by the model averaged 0.470 for the 13×13 branch, 0.404 for the 3×3 branch and 0.126 for the 33×33 branch, indicating that the two finer scales contributed most strongly on average (Fig. 2D). When expressed as the fraction of land pixels for which each branch was the largest contributor, the 13×13 scale accounted for 85.3% of pixels, followed by the 3×3 scale at 14.2%, while the 33×33 scale was dominant at only 0.5% (Fig. 2D). Pixel dominance refers to the branch receiving the largest weight at a location and does not imply that the other scales were excluded from the prediction. Regional examples from Latin America and the Caribbean, Europe, South Asia and Southeast Asia further showed that scale use was characterised by a mosaic of local and neighbourhood contexts, with broader-scale dominance forming localized concentrations, particularly in Southeast Asia (Fig. 2C). At the 5-km grid resolution used here, the 3×3 branch corresponds to an approximately 15-km local window and represents the immediate surroundings within or around a city. The 13×13 branch spans approximately 65 km and is more consistent with the scale of a metropolitan area or dense city cluster. The 33×33 branch covers approximately 165 km and represents broader regional context, such as an extended urban region, coastal belt or inhabited corridor connecting multiple population centres (Fig. 2B).

**Figure 2.**
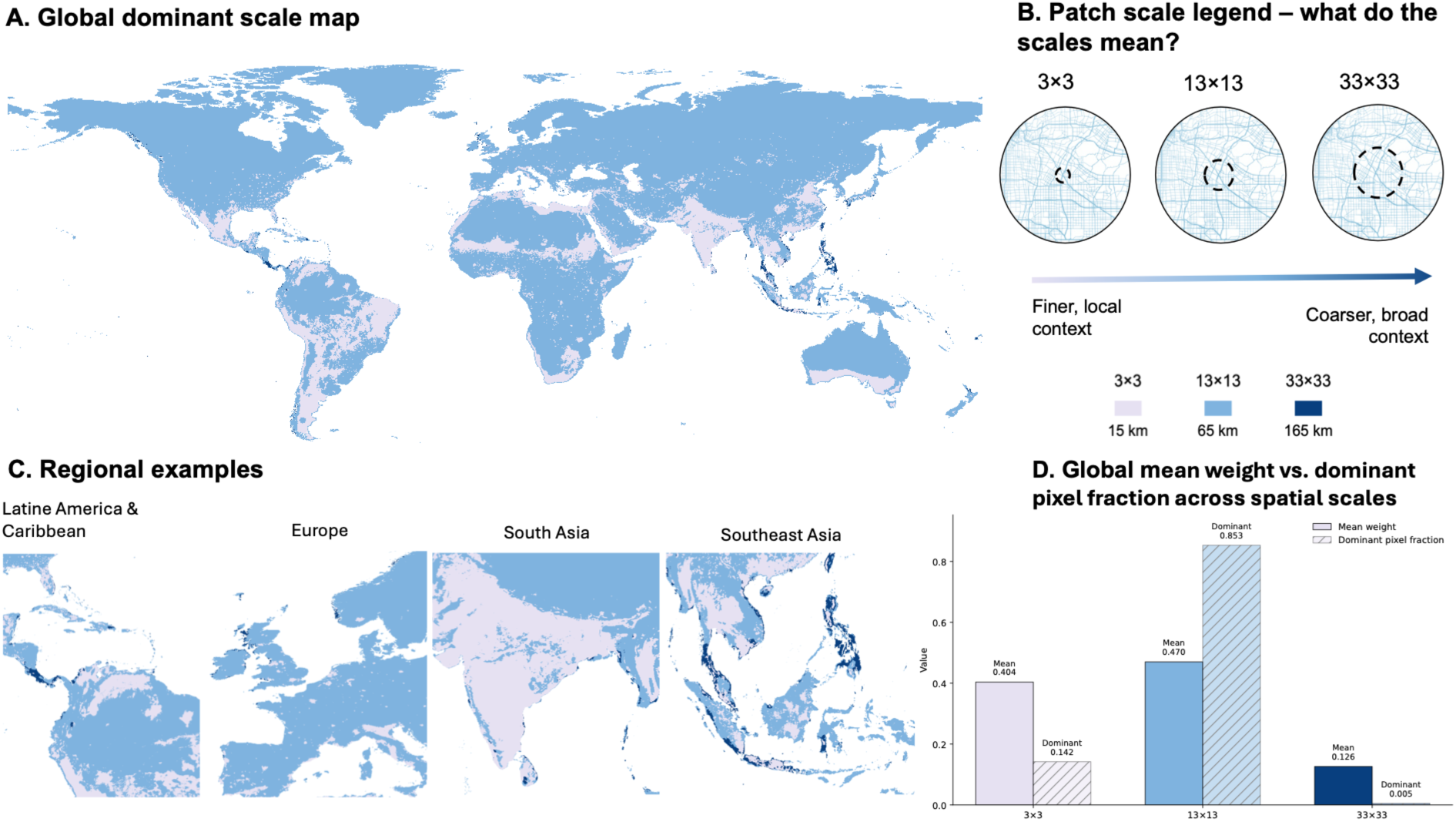
Learned spatial-scale contributions in the multiscale CNN-DRE. (A) Global map showing the dominant spatial scale at each land pixel, defined as the branch receiving the largest location-specific fusion weight. (B) Interpretation of the three patch scales: 3×3, 13×13 and 33×33 neighbourhoods, corresponding to spatial windows of approximately 15, 65 and 165 km, respectively. (C) Regional examples from Latin America and the Caribbean, Europe, South Asia and Southeast Asia showing a mosaic of local and neighbourhood-scale dominance, together with localized concentrations of broader-scale dominance, particularly in Southeast Asia. (D) Global comparison of the mean model-generated fusion weights and the fraction of pixels dominated by each scale. Mean weights were 0.404, 0.470 and 0.126 for the 3×3, 13×13 and 33×33 branches, respectively. The 13×13 branch was dominant across 85.3% of land pixels, followed by the 3×3 branch at 14.2%, while the 33×33 branch was dominant at only 0.5%. These results indicate that neighbourhood-scale context was the largest contributor across most locations, local context also contributed substantially on average, and broader context played a smaller but locally concentrated role.

The predictive contribution of each spatial scale was further assessed by comparing single-scale models, leave-one-out variants and the full multiscale CNN-DRE (Supplementary Fig. S3-D). Among the single-scale models, the 3×3 branch performed least well (Boyce = 0.888, AUC = 0.949), whereas the 13×13 and 33×33 branches achieved higher scores (Boyce = 0.933 and 0.945; AUC = 0.964 and 0.962, respectively). Compared with the 13×13 and 33×33 single-scale models, the full multiscale CNN-

DRE increased the Boyce index from 0.933 and 0.945, respectively, to 0.971, while AUC increased more modestly from 0.964 and 0.962 to 0.976. The benefit of multiscale fusion was therefore clearest for Boyce, which is particularly relevant for suitability mapping because it evaluates whether observed presences become increasingly concentrated in areas assigned higher suitability, whereas AUC primarily measures discrimination between presences and selected background locations. Removing individual scales reduced the Boyce index to 0.921 without the 3×3 branch, 0.947 without the 13×13 branch and 0.9072 without the 33×33 branch. In contrast, AUC varied only slightly across these configurations (0.975–0.976). The particularly large Boyce decline after removing the 33×33 branch indicates that broader spatial context provided complementary information despite being dominant at relatively few locations.

SHAP attribution showed that predictions from the multiscale CNN-DRE reflected a combination of demographic, socioeconomic, climatic, vector-related and ecological factors. Population density had the strongest overall contribution, followed by dengue temperature suitability, poverty index and minimum temperature of the coldest month, while enhanced vegetation index and the remaining predictors had smaller global effects (Fig. 3C). The spatial attribution map showed that the importance of these leading predictors varied geographically, with population density contributing widely across densely populated regions, poverty index becoming more prominent across parts of sub-Saharan Africa, and the two temperature-related predictors showing more regionally concentrated patterns (Fig. 3A). Regional profiles confirmed that population density was an important contributor across the Caribbean and South America, South Asia, Southeast Asia and East Africa, while secondary drivers differed among regions, particularly through stronger poverty-related contributions in Southeast Asia and East Africa and stronger thermal contributions in the Caribbean and South America and South Asia (Fig. 3B). The response curves showed a strong increase in attribution with population density, particularly at its highest values, and a general decline with increasing poverty index. Minimum temperature of the coldest month shifted from negative to positive attribution as temperatures increased, whereas dengue temperature suitability showed a more variable, nonlinear response with greater uncertainty and stronger positive attribution at its upper end (Fig. 3D).

**Figure 3.**
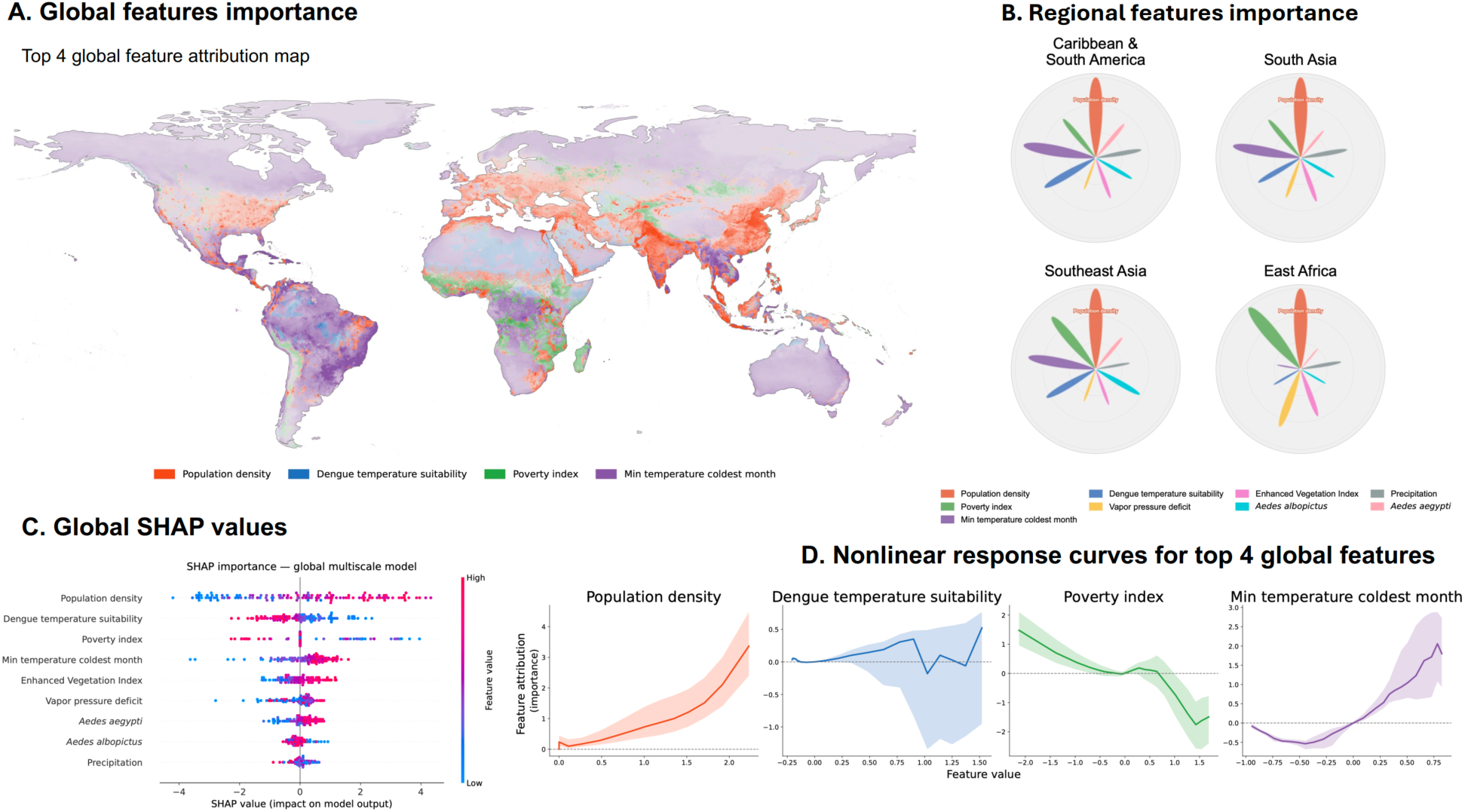
SHAP-based feature attribution for multiscale CNN-DRE dengue suitability predictions. (A) Global map of the dominant attribution among the four leading predictors: population density, dengue temperature suitability, poverty index and minimum temperature of the coldest month. (B) Regional feature-importance profiles for the Caribbean and South America, South Asia, Southeast Asia and East Africa. (C) Global SHAP summary showing population density as the strongest predictor, followed by dengue temperature suitability, poverty index and minimum temperature of the coldest month. (D) SHAP response curves showing nonlinear relationships between the four leading predictors and their model attributions. Overall, population density was the main global contributor, while the importance of secondary socioeconomic and environmental predictors varied regionally.

## 3 Discussion

This study shows that mapping global dengue suitability can be improved when locations are considered within their surrounding socio-environmental landscape rather than in isolation. By combining density-ratio estimation with multiscale spatial context, the multiscale CNN-DRE framework improved predictive performance, altered the inferred geography of suitability, and revealed the spatial scales and covariates that are most influential in model predictions.

Our results indicate that dengue suitability is shaped by scale-dependent spatial context and therefore cannot be fully captured from point-location covariates alone. In other words, the suitability assigned to a location depends partly on the surrounding configuration of population, socioeconomic and environmental conditions. Consistent with this interpretation, the multiscale CNN-DRE outperformed the point-based baselines under both target-group and uniform-background sampling, while shuffling spatial positions within patches reduced performance despite preserving their covariate content (Fig. 1 and Supplementary Fig. S1). This suggests that the improvement arose partly from spatial arrangement rather than simply from access to additional covariate values. Incorporating spatial context also did not expand predictions uniformly: the estimated at-risk surface increased in South America, East Africa, South Asia and Southeast Asia but decreased in West Africa, indicating that contextual information modified suitability in geographically specific ways. The scale-attribution analysis further showed that this effect was not simply the result of broad regional smoothing. The model relied primarily on neighbourhood and local context, while the broader scale formed localized concentrations and contributed complementary information despite rarely being dominant (Fig. 2). This interpretation was reinforced by the ablation analysis: no single-scale model matched the full multiscale CNN-DRE, and removing the broadest branch produced the largest decline in Boyce performance (Supplementary Fig. S5D). Taken together, these findings suggest that dengue suitability is most strongly structured at local-to-metropolitan scales, where human settlement and environmental conditions are likely to interact, while broader regional context provides additional information at selected locations.

The value of spatial context lies not simply in providing the model with more covariate values, but in capturing how surrounding conditions are spatially configured. Point-based models treat each grid cell as an independent covariate vector; consequently, locations with similar focal conditions may receive similar predictions even when one is embedded within a connected urban corridor and the other is geographically isolated. In contrast, the multiscale CNN-DRE can distinguish such settings through the surrounding arrangement of population, socioeconomic and environmental conditions. The shuffled-patch control supports this interpretation because destroying spatial arrangement while preserving covariate content reduced performance (Supplementary Fig. S1B). Spatial configuration therefore provided information beyond that contained in the focal values or the patch-wide covariate distributions alone.

The scale analyses also show that increasing patch size does not automatically improve suitability prediction (Supplementary Fig. S5D, Supplementary Fig. S6A). Performance varied non-monotonically across fixed patch sizes, with the 13×13 and 33×33 models performing best on different metrics, whereas combining scales produced the strongest Boyce performance (Supplementary Fig. S5-D). Patch size should therefore be treated as an ecological design choice rather than simply as a technical hyperparameter. At the 5-km resolution used here, the 3×3, 13×13 and 33×33 patches represent approximately 15, 65 and 165-km windows, corresponding broadly to local, metropolitan and regional context. These distances should not be interpreted as direct transmission radii; instead, they describe the geographic extent over which surrounding covariate patterns informed the prediction. The same 13×13 patch applied to 1-km rasters would span only 13 km and represent a substantially different ecological context. Future applications should therefore define patch sizes in geographic units and evaluate windows spanning the expected local, neighbourhood and broader scales of the processes under study.

Feature-attribution analyses suggest that the multiscale CNN-DRE learned a socio-environmental signature of dengue suitability rather than a purely climatic envelope. Population density was the strongest global predictor, followed by dengue temperature suitability, poverty index and minimum temperature of the coldest month (Fig. 3C). The strong positive attribution at high population densities is consistent with the importance of concentrated human settlement for sustaining dengue transmission. However, this population signal was modified by different secondary conditions across regions. Poverty-related contributions were more prominent in Southeast Asia and East Africa, whereas temperature-related predictors contributed more strongly in the Caribbean and South America and in South Asia (Fig. 3A,B). The nonlinear response curves further indicated that these associations were not uniform: population-density attribution increased sharply at high values, minimum temperature of the coldest month became increasingly positive under warmer conditions, poverty attribution generally declined as the index increased, and dengue temperature suitability showed a more variable and uncertain response (Fig. 3D). These regional differences suggest that similar levels of predicted suitability can arise from different combinations of demographic, socioeconomic and environmental conditions.

The scale-specific SHAP summaries add a second layer to this interpretation. At the local 3×3 scale, population density and poverty index were the two leading predictors, indicating a strong contribution from immediate human and socioeconomic context. At the 13×13 neighbourhood scale, dengue temperature suitability became the leading predictor, followed by population density, minimum temperature of the coldest month, vegetation and vapour-pressure deficit. At the broader 33×33 scale, temperature-related predictors remained prominent, while Aedes albopictus suitability became more influential and poverty index contributed little (Supplementary Fig. S5). This shift suggests that the different branches learned complementary information: local predictions were more strongly structured by population and socioeconomic gradients, whereas neighbourhood and regional branches placed greater emphasis on climatic, ecological and vector-related conditions. For example, a highly deprived but sparsely populated rural location may receive a different attribution pattern from a dense urban or peri-urban location even when both have similar poverty values. The negative poverty attribution should therefore be interpreted as a conditional model association after accounting for correlated predictors, not as evidence that poverty is protective or causally reduces dengue suitability.

The geographically varying suitability patterns identified by the multiscale CNN-DRE have practical implications for dengue surveillance and preparedness. By identifying areas of suitability that differed from those predicted by point-based baselines, the model may highlight populations living in environments compatible with dengue occurrence but underrepresented in existing occurrence records. This is particularly relevant for regional margins and transition zones, such as peri-urban belts around large cities, connected settlement corridors, coastal or riverine population clusters, and areas bordering known endemic zones where local covariates alone may not capture the surrounding urban, ecological or climatic context. These areas could be useful targets for strengthened entomological surveillance, diagnostic capacity and early-warning systems. From a policy perspective, these findings support combining targeted local interventions with coordinated surveillance and preparedness across metropolitan and regional scales, rather than treating at-risk locations as isolated units. However, the population-at-risk estimates should be interpreted as populations living in areas of predicted suitability, not as expected dengue incidence. Suitability maps identify where dengue occurrence is environmentally and socio-demographically plausible; they do not directly account for immunity, serotype dynamics, vector-control interventions, healthcare access, reporting effort or short-term outbreak conditions. More broadly, the framework is not dengue-specific and could be adapted to other vector-borne or environmentally mediated diseases whose occurrence depends on both local conditions and neighbourhood-scale landscape context.

## 4 Limitations

Several limitations should guide interpretation of these results. First, CNN-DRE estimates relative suitability with respect to the background distribution used during training. In this study, background points were sampled using a target-group strategy designed to approximate the sampling bias of the presence data. This reduces the contrast between well-sampled presence locations and unrealistic or weakly sampled background environments, such as sparsely inhabited areas where sustained dengue transmission is unlikely. However, it also means that the fitted density ratio should not be interpreted as an absolute measure of suitability over all possible environments. Instead, predictions are relative to the constrained background against which presences were compared. Careful design of the background distribution therefore remains central to both model validity and interpretation.

A second limitation is the absence of an explicit temporal component. Although spatio-temporal modelling could in principle better represent seasonal and interannual variation in dengue transmission, the occurrence data used here were temporally heterogeneous, with variation likely reflecting changes in surveillance, reporting, and data availability as much as ecological dynamics. Introducing time directly under these conditions could therefore risk modelling reporting history rather than transmission processes. Our model should instead be interpreted as estimating structural spatial suitability, not short-term outbreak risk or temporal variation in incidence. Spatio-temporal extensions remain an important direction for future work, but will require occurrence data with more consistent temporal coverage and better control for surveillance effort.

The modelling framework also represents a deliberate trade-off between flexibility, interpretability, and feasibility. More expressive density-ratio estimators, including latent-space or generative approaches such as normalizing flows^24^, could in principle capture more complex distributions. However, these models are typically more data-demanding, computationally intensive, and harder to interpret. This is a non-trivial constraint in disease distribution modelling, where presence data are often limited, spatially biased, and intended to inform public-health decisions. Our model increases expressiveness relative to point-based baselines while remaining lightweight enough to train with modest computational resources and amenable to post-hoc interpretation through scale attribution and SHAP analyses.

Finally, we did not estimate dengue detection probability directly. Instead, Zika and chikungunya occurrence records were used to construct a target-group background representing the spatial distribution of surveillance and reporting for related Aedes-borne arboviruses. Within the density-ratio formulation, spatial sampling processes common to dengue presences and the target-group background are attenuated because they occur in both the numerator and denominator. This approach therefore reduces bias associated with shared spatial variation in sampling effort, while acknowledging that residual bias may remain where dengue-specific detection or reporting differs from that of the target group.

## 5 Conclusion

In conclusion, this study shows that global dengue suitability is better represented when locations are modelled within their surrounding socio-environmental context rather than as isolated points. By combining density-ratio estimation with mask-aware multiscale CNNs, the multiscale CNN-DRE improved predictive performance, produced more spatially coherent suitability maps, and identified additional populations and areas living in environments compatible with dengue occurrence. The learned scale and feature attributions further showed that this improvement was driven mainly by local-to-neighbourhood spatial structure and a dominant human-settlement signal, with regional modulation by climatic, ecological, vector-related, and socioeconomic factors. More broadly, the framework provides an adaptable approach for mapping environmentally structured disease risk from biased occurrence data, while retaining interpretability through scale attribution, SHAP analysis, and explicit evaluation of modelling choices.

## 6 Methodology

### 6.1 Data and preprocessing

#### 6.1.1 Data

##### Virus occurrences

Species distribution models require both presence data, indicating where a species has been observed, and absence data, indicating where it does not occur, to relate species occurrence to environmental conditions. True absence of disease is difficult to ascertain due to global heterogeneities in disease surveillance capacity. Here, we used a presence-only dataset of georeferenced human dengue cases^8^, which were originally collected from various sources including: Messina et al. (2019)^7^, the Diseases Outbreak News^25^ and HealthMap platforms^26^. Occurrence records were highly uneven in temporal coverage. As shown in Supplementary Figure S7, reporting increased markedly after 2000, with most observations concentrated in recent years, likely reflecting changes in surveillance and reporting practices rather than purely ecological dynamics. Reporting was also spatially heterogeneous: 36,750 records corresponded to 8,135 unique locations, with a small subset of sites accounting for a disproportionate share of observations. Under these conditions, spatio-temporal modelling would primarily capture variation in reporting effort rather than transmission dynamics. We therefore focused on estimating spatial suitability based on locations where dengue transmission has been documented at least once during 2010-2023. Reporting dates were not incorporated, and records were deduplicated by location. The model thus characterizes structural environmental suitability rather than temporal variation in incidence.

##### Background points

A key challenge in disease distribution modelling is the lack of true absence data. This is commonly addressed through background or pseudo-absence sampling. Background points represent the environmental space available within the study area and do not imply presence or absence, whereas pseudo-absences are treated as absences despite the lack of confirmation. In this study, we use background points as a reference against which the environmental characteristics of presence locations are contrasted for model training and evaluation. Several approaches are used to sample background points, including uniform random sampling across the study area, environmentally stratified sampling, and target-group (similar species) background sampling^27,28^. The latter has strong empirical support for reducing sampling bias in species distribution models^29,30^. In target-group sampling, background points are drawn from the same sampling process as the presence data, ensuring that background and observations share the same bias structure. Recent work has further refined this approach. For example, Poongavanan et al. (2025)^31^ demonstrated that applying a target-group background with a spatial buffer (50-500 km) around presence locations, combined with kernel density estimation (KDE) smoothing (bandwidth = 5), enhances predictive performance in top-performing SDM algorithms such as RF. These methodological refinements reduce both spatial sampling bias and artificial inflation of model accuracy.

Building on these considerations, we adopted a target-group background approach, using Zika and chikungunya occurrence records from Lim et al. (2025)^8^. These arboviruses share the primary vectors *Aedes aegypti* and *Aedes albopictus* with dengue and are subject to similar surveillance systems, thereby approximating comparable sampling biases. Background points were sampled within a 50-500 km buffer around presence records, with KDE based thinning applied to approximate underlying sampling effort and mitigate spatial clustering.

##### Covariates

The spatial distribution of dengue and its vectors is shaped by interacting environmental, climatic, and socio-economic determinants. Covariate selection for the model was guided by the systematic review by Lim et al. (2023)^16^, which synthesizes evidence on determinants of Aedes-borne arbovirus transmission. All covariates were compiled for the period 2010-2023. Environmental and climatic variables included precipitation^32^, Enhanced Vegetation Index (EVI)^33^, Vapor Pressure Deficit (VPD)^32^ and minimum temperature of the coldest month^32^. Socio-economic conditions were represented by population density^34^ and a poverty/deprivation index^35^, capturing variation in human exposure and vector-host contact intensity. In addition, we included model-derived suitability layers as spatial covariates: global distribution maps of *Aedes aegypti* and *Aedes albopictus* from Kraemer et al. (2015)^5^, and a temperature-based dengue transmission suitability index from Brady et al. (2014)^6^. These outputs provide validated, spatially explicit summaries of vector presence and temperature-driven suitability and were incorporated alongside the primary predictors.

#### 6.1.2 Preprocessing

##### Standardisation and normalization

After covariate selection, all raster layers were standardised and reprojected to EPSG:4326 on a common global grid to ensure spatial alignment. Using the precipitation layer as the reference, all rasters were resampled to a uniform 5 km resolution and clipped to a common spatial extent. Feature scaling is known to influence optimisation behaviour and predictive performance in machine learning models^36^. Prior to training, all standardised raster covariates were individually normalised using a Median-IQR transformation also known as Robust scaler, which centres values on the median and scales them by the interquartile range. This corresponds to a robust estimate of location and scale and is less sensitive to extreme values than normalisation based on the sample mean and standard deviation^37^. Applying this transformation places all covariates on comparable scales while limiting the influence of extreme values, which is particularly appropriate for environmental data subject to heterogeneous measurement conditions (supplementary Fig. S9).

##### Patch extraction

Patches form the core spatial inputs to our model and provide local spatial context alongside multivariate environmental information. Ecological suitability often depends on surrounding conditions such as habitat configuration, fragmentation, and edge effects rather than on single pixels, because many ecological processes operate at neighbourhood scales^38^. Patch-based inputs therefore allow convolutional neural networks to exploit their receptive fields to learn spatial dependencies and nonlinear interactions among environmental variables and their local spatial arrangement^20,22^.

We extracted small, fixed-size image patches centred on each sampled location for both presence and background points. Each patch represents a local environmental “window” that captures ecologically meaningful spatial patterns in the neighbourhood surrounding a site; patterns that cannot be represented by single-pixel values alone. Each patch is paired with a binary observation mask of the same size (Supplementary Fig. S10), in which pixels with observed values are assigned a value of 1 and pixels with missing values are assigned a value of 0.

Patch size was a key design parameter because it determined the spatial extent of the contextual information available to the model. We first conducted a fixed-scale sensitivity analysis using patch sizes of 3 × 3, 13 × 13, 23 × 23, 33 × 33 and 63 × 63 pixels. Each patch size was evaluated separately, while all other preprocessing steps and model hyperparameters were held constant to isolate the effect of patch size. We then extended this fixed-scale approach in the final multiscale CNN-DRE by simultaneously incorporating patches of 3 × 3, 13 × 13 and 33 × 33 sizes. These scales captured fine local conditions, intermediate neighbourhood patterns and broader spatial context, respectively. This multiscale design allowed the model to integrate information across spatial scales within a single architecture and learn the relative contribution of each scale to the prediction at each location.

### 6.2 Model

#### 6.2.1 Problem formulation

Our goal is to build a contemporary global suitability map for dengue virus at 5km resolution. Specifically, for any geographic location, we aim to estimate the probability of dengue occurrence given climatic, environmental and socio-economic conditions extracted from raster data.

Each location is represented by a multi-covariate raster patch:

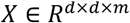

where *d* represents the spatial dimension of the patch and *m* the number of raster layers (climatic, ecological, demographic, etc.). Thus, *X* encodes both spatial context and multiple covariates.

Let Y ∈ {0,1} denote the presence (Y = 1) or absence (Y = 0) of Dengue.

Our target is the conditional probability:

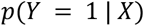

Using Bayes’ theorem:

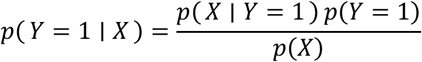

Here, p(X ∣ Y = 1) is the distribution of raster patches at confirmed presence locations, while p(X) denotes the distribution of patches under our chosen background sampling scheme. The factor p(Y = 1) reflects the overall rate of dengue occurrence, which is not identifiable from presence-background data and cannot be estimated reliably at the global scale. As a result, absolute probabilities p(Y = 1 ∣ X) are not estimable.

Instead, we focus on relative suitability, defined as:

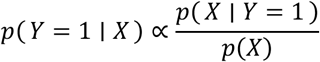

This quantity measures how likely a given set of environmental conditions occurs at presence locations relative to the background landscape. Estimating it is a density-ratio estimation problem, which we present in the next section. Specifically, it is the ratio of the density of environmental conditions at presence locations to their density at background locations. For a specified background distribution, the resulting relative-suitability surface provides an identifiable characterization of dengue’s ecological niche, even when true prevalence and absolute occurrence probabilities are unavailable.

#### 6.2.2 Model description

Supplementary Figure S3 provides an overview of the CNN-DRE architecture and evaluation workflow. Panel (i) presents the single-scale CNN-DRE, in which a mask-aware CNN encodes masked socio-environmental raster patches and an MLP-based density-ratio estimation (DRE) head maps the resulting representations to relative suitability scores. Panel (ii) outlines the spatial clustering, cross-validation, model-selection, and held-out test-evaluation pipeline used throughout the study. Panel (iii) presents the multiscale extension, which extracts contextual information at three nested spatial extents (3 × 3, 13 × 13, and 33 × 33), combines the resulting representations using an adaptive fusion module, and passes the fused representation to the same DRE head. The following sections first describe the core CNN-DRE components and then introduce the multiscale extension.

##### Encoder

An encoder *f*_$_ is employed to transform patches into compact feature vectors:

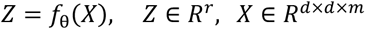

Where *Z* is an *r*-dimensional representation of summarising local contextual information for density-ratio estimation.

The encoder is a CNN, meaning it applies small learnable filters repeatedly across the patch to detect spatial patterns that may occur anywhere within the neighbourhood. By stacking several such layers, the encoder can represent increasingly complex local structure from simple spatial contrasts in the covariates to more integrated contextual signatures while keeping the number of parameters manageable through weight sharing.

A central challenge in gridded environmental covariates is the presence of missing or invalid pixels (e.g., outside the study extent or no-data regions). Standard convolutions treat missing values as ordinary numbers (often zeros), which can introduce artefacts and allow the model to learn from missingness rather than environmental structure. To address this, we use partial convolutions^39^, which incorporate a binary validity mask *M* and compute each local convolution using only observed pixels. Concretely, within each *k* × *k* neighbourhood (here *k* = 3), pixels flagged as invalid by *M* are ignored, meaning their contribution is set to zero in the local sum, and the convolution output is then re-scaled by the number of valid pixels in that neighbourhood so that responses remain comparable across locations with different amounts of missing data. If no valid pixels are available in a neighbourhood, the output at that location is set to zero and the location remains marked invalid for subsequent layers. We apply a channel wise mask in the first layer, allowing missingness to differ across covariate channels and then propagate a single spatial mask through the remainder of the encoder.

Architecturally, *f_θ_* is composed of a stack of 3 × 3 partial-convolution blocks, each followed by batch normalization to stabilise feature magnitudes during training and a Rectified Linear Unit (ReLU) for nonlinearity. The same general encoder design was used across scales, with pooling depth adjusted to the spatial extent of the input: the 3 × 3, 13 × 13, and 33 × 33 branches used zero, one, and two mask-aware pooling operations, respectively, thereby avoiding excessive spatial reduction at the smallest scale while providing efficient context aggregation at larger scales. The validity mask was pooled in parallel, such that a pooled location was considered valid if any contributing pixel was valid. Where appropriate for the available spatial extent, intermediate blocks used dilated partial convolutions to expand spatial context without additional downsampling. Finally, each encoder aggregated its feature maps using mask-aware global average pooling, which averages only over valid spatial locations, and mapped the resulting vector through a linear projection to produce the final embedding *Z* (with r=32 in our experiments). These scale-specific encoder configurations were retained unchanged in the sensitivity analysis.

##### Multiscale CNN-DRE

The multiscale CNN-DRE extends the general CNN-DRE framework by replacing the single fixed-scale patch encoder with three parallel mask-aware CNN branches operating at nested spatial extents. For each prediction location, we used three nested spatial windows of 3×3, 13×13 and 33×33 pixels, representing fine local, neighbourhood and broader surrounding context, respectively. Each branch receives both the covariate values and their associated validity masks, and processes them with the same mask-aware partial-convolutional encoder described above. In this way, the model learns three scale-specific embeddings, each summarising the socio-environmental context around the focal location at a different spatial extent.

The three embeddings are combined through an adaptive fusion module whose parameters are shared across the study area but which generates input-dependent weights separately for each prediction location. The weighted embeddings are concatenated into a single fused representation and passed to the same MLP-based density-ratio head used in the general CNN-DRE architecture. This allows the relative contribution of each spatial scale to vary according to the characteristics of the focal location rather than imposing a fixed spatial extent everywhere. For interpretation, we reported global average weights by summarising these location-specific weights across the study area. A study-area-wide map of locally dominant scale was also produced by assigning each prediction location to the branch receiving the largest adaptive fusion weight.

##### Multi-Layer Perceptron (MLP) Density Ratio Estimation head

Density-ratio estimation (DRE) refers to methods that directly estimate 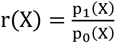 without separately estimating the densities p_1_ and p_O_ which in our case respectively represent presence and background densities. In this work, we implement four classic framework for DRE, developed by Sugiyama et al. (2010, 2012)^40^: probabilistic classification, Kullback-Leibler Importance Estimation Procedure (KLIEP), unconstrained Least-Squares Importance Fitting (uLSIF) and Kernel uLSIF (KuLSIF) via a Multi-Layer Perceptron (MLP).

A MLP is a feed-forward neural network composed of a sequence of fully connected layers that apply nonlinear transformations to an input vector. In this study, the MLP is used to map the latent representation produced by the convolution encoder to a single scalar score representing relative environmental suitability. Specifically, the CNN encoder outputs a low dimensional embedding vector Z ∈ R^r^ for each environmental patch, summarising local environmental conditions in a fixed-length representation. This vector is then provided as input to the MLP, denoted g**_φ_**. The MLP consists of a sequence of fully connected layers with nonlinear activations and dropout regularization, followed by a final linear output layer. The dropout regularization is applied during training to reduce overfitting and improve generalization by randomly deactivating a fraction of hidden units (neurons.

Given an embedding Z, the head outputs a scalar logit s = g**_φ_**(Z), which parameterizes the density ratio between two distributions of interest: the latent representation of presence locations and those of background locations. The role of this network is not features extraction, but rather the flexible approximation of a scalar function defined on the learned latent space. All the four DRE techniques we implemented rely on the same MLP architecture but differing in the training objective used to estimate the density ratio between presence and background distributions in the latent space.

##### Probabilistic classification (classifier-based DRE)

In this approach, density-ratio estimation is recast as a binary classification problem between samples drawn from the presence distribution P_1_(Z) and the background distribution P_O_(Z). The MLP outputs the score s = g**_φ_**(Z), which is trained using the binary cross-entropy loss. At optimality, the classifier estimates the posterior probability

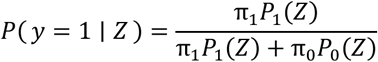

where π_1_ and π_O_ denote class priors. The density ratio is then recovered analytically as

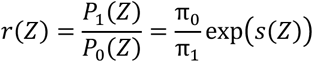

To train the classifier, we use binary cross-entropy with logits (BCE). Here, *s*(*Z*) is a logit (an unbounded real-valued score), and the corresponding probability of presence is obtained through the logistic sigmoid function 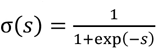. For a sample with label *y* ∈ {0,1}, the BCE loss is:

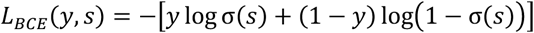

This loss decreases when presence samples (y = 1) receive high scores and background samples (y = 0) receive low scores, thereby learning a discriminative score function in latent space.

In addition to BCE, we include a pairwise ranking loss to improve the ordering of predictions. This is useful because discrimination metrics such as ROC AUC depend primarily on the relative ranking of scores rather than on probability calibration. For a presence sample i and a background sample j, with logits *s_i_* and *s*_j_, the ranking loss uses the score difference *s_i_* − *s*_j_and penalizes cases where this difference is too small or negative:

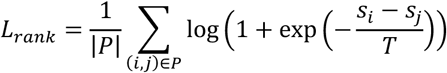

where P is the set of presence-background pairs in a mini-batch.

The parameter *T* > 0 is a temperature, which controls the smoothness/sensitivity of the ranking penalty: smaller *T* makes the loss more sensitive to score differences (sharper penalty), while larger *T* softens the penalty and reduces gradient magnitude. In our implementation, *T* = 1.

The final training objective is a weighted sum of the classification and ranking terms:

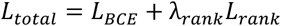

where λ_rank_ controls the contribution of the ranking term. This combined objective encourages both accurate presence/background discrimination and good score ordering for downstream density-ratio-based suitability ranking.

For the comparison of classifier-based DRE with KLIEP, uLSIF, and KuLSIF, only the standard BCE formulation was used. Because classifier-based DRE achieved the best comparative performance, it was retained as the basis of the final model, in which the auxiliary ranking loss and AUC-weighted ensembling of fold logits were introduced to improve discrimination and predictive stability. As these adaptations prioritise ranking and robustness rather than strict probability calibration, the final ensemble output is interpreted as a relative density-ratio score rather than an exactly calibrated estimate of the absolute density ratio.

##### Kullback-Leibler Importance Estimation Procedure (KLIEP)

KLIEP directly models the density ratio r**_φ_**(Z) without estimating either density. In this case, the MLP is trained to maximize

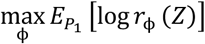

subject to the normalization constraint

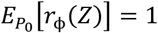

In this setting, the MLP output is interpreted as log r**_φ_** (Z), and the constraint can be enforced explicitly or via a penalty term.

The MLP output is interpreted as the log-density ratio, i.e. ℎ**_φ_**(*Z*) = log *r***_φ_**(*Z*) and the ratio is parameterized as *r***_φ_**(*Z*) = exp Yℎ**_φ_**(*Z*)[, which guarantees positivity. In our deep KLIEP implementation, the MLP therefore learns a scalar scoring function ℎ**_φ_**(*Z*) on the latent embedding space, where larger values indicate embeddings that are more characteristic of presence samples than background samples.

Using this parameterization, the KLIEP objective can be written as the minimization of the following loss:

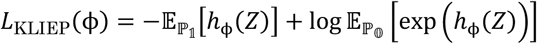

The first term encourages the model to assign high scores to presence embeddings, while the second term enforces normalization with respect to the background distribution and prevents trivial solutions in which the ratio is increased everywhere. Here, D_P1_[⋅] and D_P0_[⋅] denote expectations (approximated in practice by minibatch averages) over presence and background embeddings, respectively. Intuitively, deep KLIEP learns a direct estimate of relative environmental suitability by assigning large density-ratio values to regions of the latent space that are common under the presence distribution and rare under the background distribution.

##### Unconstrained Least-Squares Importance Fitting (uLSIF)

uLSIF estimates the density ratio by minimizing the squared-error objective

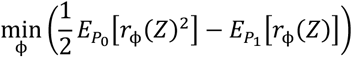

Equivalently (up to an additive constant independent of **φ**), this objective corresponds to minimizing the squared error

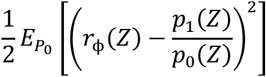

which shows that the optimum recovers the true density ratio when the function class is sufficiently expressive. In practice, the expectations are replaced by empirical minibatch averages over samples from the two groups (y = 0 for ***P*_0_**, y = 1 for *P*_1_), yielding the training loss:

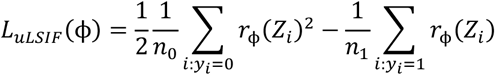

This is called unconstrained because it does not impose an explicit normalization constraint on the ratio model (e.g. ∫ *r*(*z*)*p*_O_(*z*) *dz* = 1) during optimization. Instead, the ratio is learned directly by minimizing the squared-error surrogate. In our implementation, *r***_φ_**(⋅) is represented by the MLP, and a nonnegative output activation is used so that the estimated ratio remains physically meaningful (*r***_φ_**(*Z*) ≥ 0).

Here, the density ratio also called an importance ratio quantifies how much more likely a feature vector Z is under the target distribution *P*_1_ than under the reference distribution *P*_O_. The expectations D_P0_[⋅] and D_P1_[⋅] denote averages with respect to samples drawn from these two distributions.

##### Kernelized uLSIF (KuLSIF)

KuLSIF extends uLSIF by adding explicit regularization to control model complexity and improve robustness. In practice, the quantity minimized during training is the loss function:

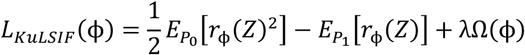

Where λ > 0 is a regularization parameter. As in previous cases, *r***_φ_**(*Z*) is the density-ratio estimator that aims to approximate r(Z). The loss works by balancing two effects: 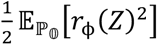 penalizes overly large ratio values on background samples, while 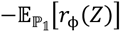 encourages larger ratio values on target samples. The regularization term λ Ω(**φ**) controls model complexity, improves numerical stability, and reduces overfitting.

Out-of-fold model-development analyses were used to select the DRE objective, spatial scales and loss configuration (Fig. 4A,B). Across patch sizes, classifier-based DRE provided the most stable balance between Boyce performance and training time, whereas uLSIF and KuLSIF deteriorated at larger spatial windows and KuLSIF incurred the greatest computational cost (Supplementary figure S6.A). Performance did not increase monotonically with patch size, with the classifier reaching its strongest fixed-scale performance at 13×13 and 23×23. Within the classifier-based approach, adding the ranking term improved Boyce performance at all evaluated scales, with the largest gains at 13×13, from 0.912 to 0.9645, and 33 × 33, from 0.876 to 0.926 (Supplementary figure S6.B). These analyses supported the use of classifier-based DRE with the combined BCE and ranking objective in the final multiscale CNN-DRE.

#### 6.2.3 Model Evaluation

We adopted a spatially explicit cross-validation strategy designed to minimize geographic bias due to spatial autocorrelation and to evaluate true out-of-sample generalization. Occurrence locations are first embedded on the Earth’s sphere using three-dimensional unit vectors and partitioned with k-means clustering, yielding a fixed total of 50 spatially coherent clusters of presence points. Background points are subsequently assigned to these clusters by nearest-centroid prediction. Clusters, rather than individual points, constitute the atomic units of data splitting, ensuring that geographically proximate observations are never separated across training and validation sets. The same spatially explicit cross-validation, held-out test design, and fold-ensemble evaluation strategy were applied to all CNN-DRE variants, including the final multiscale model.

From these 50 clusters, we randomly selected 5 clusters as a fixed external test set, corresponding to approximately 10% of the spatial partitions. These test clusters are selected prior to model fitting and are completely excluded from model training and hyperparameter tuning, thereby defining a spatially disjoint evaluation region. The remaining 45 clusters are then used for model development and are distributed across 10 cross-validation blocks or folds using a constrained balancing procedure that enforces at least one presence per fold and approximately equal background sample sizes. The final predictor is an ensemble of all fold-specific models. Each fold model is saved at its best validation AUC. At inference time, logits from each model are combined via a weighted sum, where weights are proportional to each fold’s validation AUC.

We finally evaluate model performance using the spatially structured validation protocol described above, with the continuous Boyce index as the primary criterion. The Boyce index measures whether locations with observed presences tend to receive higher predicted suitability than expected by chance, with values near 1 indicating strong agreement and values near 0 indicating random performance. Because model outputs are interpreted through a density-ratio or suitability perspective rather than as calibrated probabilities, we chose the Boyce index over purely classification-based metrics. The cross-validation training and evaluation strategies are visible in Supplementary figure S3.

#### 6.2.4 Baselines and point-based comparator

We compared our framework against two widely used approaches in species distribution modelling, MaxEnt and Random Forest (RF), in order to benchmark the predictive performance of the CNN-DRE models against established disease-distribution baselines. In addition, we evaluated a point-based DRE comparator, implemented as a 1×1 MLP-DRE, to isolate the effect of adding spatial context. All comparison models were trained and evaluated using the same cross-validation, prediction and evaluation protocol described above, ensuring a fair and controlled comparison. Both external baselines and the point-based DRE comparator relied exclusively on tabular environmental covariates extracted from the same rasters used to generate the CNN-DRE patches and used the same occurrence and background points.

MaxEnt is a presence-background species distribution modelling method that estimates relative habitat suitability from environmental predictors by finding the probability distribution with maximum entropy subject to constraints derived from observed presence locations. In our analysis, the MaxEnt baseline is implemented using the *elapid* maximum entropy model in Python^41^. The implemented model estimates species suitability as a regularized log-linear function of environmental covariates. To capture both linear and nonlinear responses, the model is fitted using combinations of feature classes, including linear, quadratic, hinge, and product terms. Model complexity is controlled via a beta multiplier that regulates the strength of regularization. Beta values in the range [0.25, 5.0] are considered, allowing for varying degrees of smoothness in the fitted response. Final predictions are produced using the logistic transformation, yielding continuous suitability scores in the interval [0, 1].

Random forest is an ensemble learning method that combines multiple decision trees trained on resampled data with feature randomness to improve generalization and reduce overfitting. Here, the RF baseline was implemented as a probabilistic ensemble of trees trained on standardized environmental covariates, with tree complexity controlled through split-size regularization rather than a fixed depth limit. Suitability predictions were obtained by averaging class probabilities across trees. Model capacity was tuned by selecting the number of trees from 50 to 1500 during cross-validation, and final test predictions were generated using an AUC-weighted ensemble of the fold-specific selected models.

The 1×1 MLP-DRE was included as a point-based comparator within the CNN-DRE framework. In this setting, the environmental input at each location is reduced to the focal pixel only, so no surrounding spatial context is provided. The model therefore bypasses the CNN encoder and uses only the MLP-based density-ratio estimation head operating on the tabular covariate vector extracted at the focal location. This comparator isolates the contribution of spatial context by retaining the same density-ratio formulation while removing neighbourhood information.

#### 6.2.5 Mapping of populations and global areas at risk for dengue

Global gridded population estimates for 2020, aggregated to approximately 1 km spatial resolution, were obtained from WorldPop^42^. Population at risk was estimated by overlaying the 2020 gridded population dataset with model-predicted dengue suitability maps. Population rasters were resampled to the spatial resolution and extent of each suitability model, and areas with predicted suitability >0.7 were classified as suitable. The population within suitable grid cells was summed to estimate the population at risk. The corresponding suitable area was calculated using cell-specific surface areas. These metrics were calculated globally and across predefined regions for each model.

## Data Availability Statement

Current climate data are available at https://github.com/carlfoka/Deep-Learning-for-Disease-Distribution-Modelling.

Data and code are available at https://github.com/carlfoka/Deep-Learning-for-Disease-Distribution-Modelling.

## Contributors

C.F.T. and H.T. conceptualised and designed the study. C.F.T. led the data analysis, developed and implemented the models and analyses, and produced the primary data visualisations with assistance from H.T and J.P. C F.T. wrote the original draft with assistance from J.P., H.T., M.U.G.K., E.S. and T.d.O.. H.T. supervised the work. H.T. and T.d.O. acquired funding for the study. All authors reviewed and approved the manuscript.

## Data Availability

All data produced are available online at
https://github.com/carlfoka/Deep-Learning-for-Disease-Distribution-Modelling

## Acknowledgements

The authors acknowledge that this work is an initiative of the CLIMADE consortium. The Centre for Epidemic Response are supported in part by grants from the Medical Research Foundation (MRF-RG-ICCH-2022-100069), the Wellcome Trust for the Global.health project (228186/Z/23/Z), and the Novo Nordisk Foundation (NNF24OC0094346), the Rockefeller Foundation (HTH 017), the INFORM Africa project through the Institute of Human Virology Nigeria (U54 TW012041), Global Health EDCTP3 Joint Undertaking and its members and the Bill & Melinda Gates Foundation (101103171), and the Health Emergency Preparedness and Response Umbrella Program, managed by the World Bank Group (TF0B8412). MUGK acknowledges funding from The Rockefeller Foundation (PC-2022-POP-005), Health AI Programme from Google.org, the Oxford Martin School Programmes in Pandemic Genomics & Digital Pandemic Preparedness, EU Horizon Europe programme project MOOD (874850), E4Warning (101086640), the Wellcome Trust (225288/Z/22/Z, 226052/Z/22/Z, and 228186/Z/23/Z), UK Research and Innovation (APP8583), and the Medical Research Foundation (MRF-RG-ICCH-2022-100069); UK International Development (301542-403), the Bill & Melinda Gates Foundation grants (INV-063472, INV-090281). The content and findings reported herein are the sole deductions, views, and responsibility of the researchers and do not necessarily reflect the official position and sentiments of the funding agencies.

## Supplementary Information

Supplementary Information is available for this paper.

## Correspondence and requests for materials

Correspondence and requests for materials should be addressed to Houriiyah Tegally.

## Supplementary Material

**Supplementary Figure S1.**
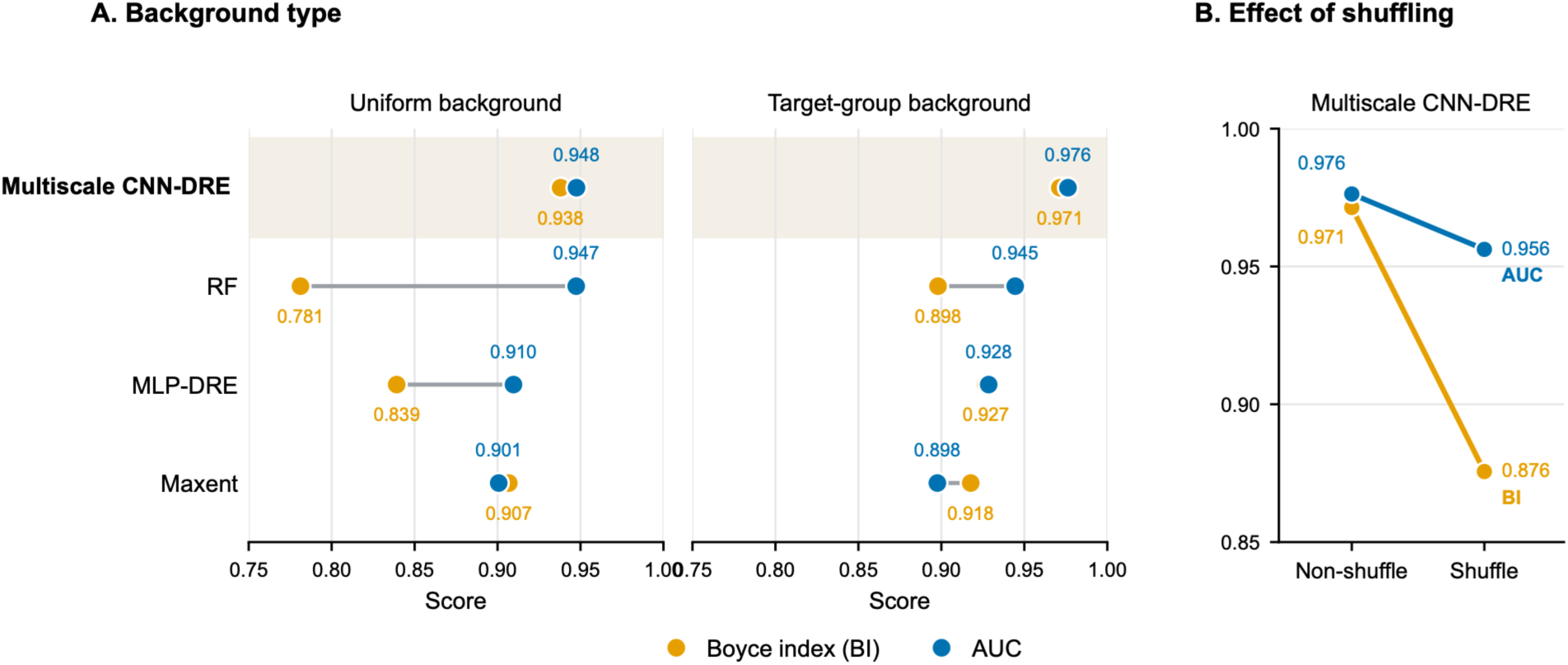

**Supplementary Figure S2.**
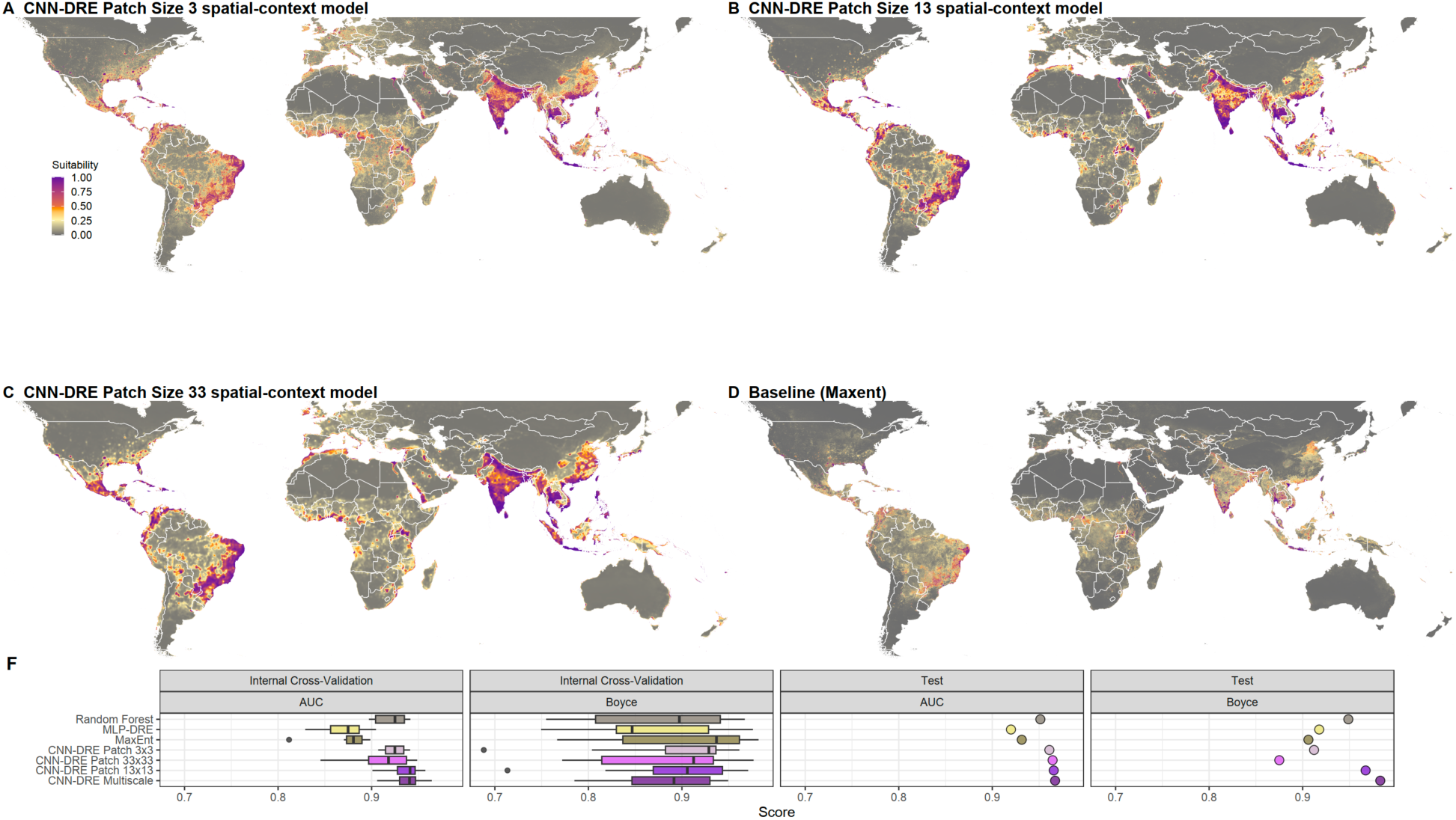
Fixed-scale CNN-DRE suitability maps and model performance comparison. (A-C) Global dengue suitability predictions from single-scale CNN-DRE spatial-context models using 3×3, 13×13, and 33×33 input patches. (D) Baseline suitability prediction from MaxEnt. (F) Model performance comparison across internal cross-validation and held-out test evaluation using AUC and Boyce index. Results compare the multiscale CNN-DRE model, fixed-scale CNN-DRE variants, and point-based baselines, showing how prediction patterns and performance vary with spatial context scale.

**Supplementary Figure S3.**
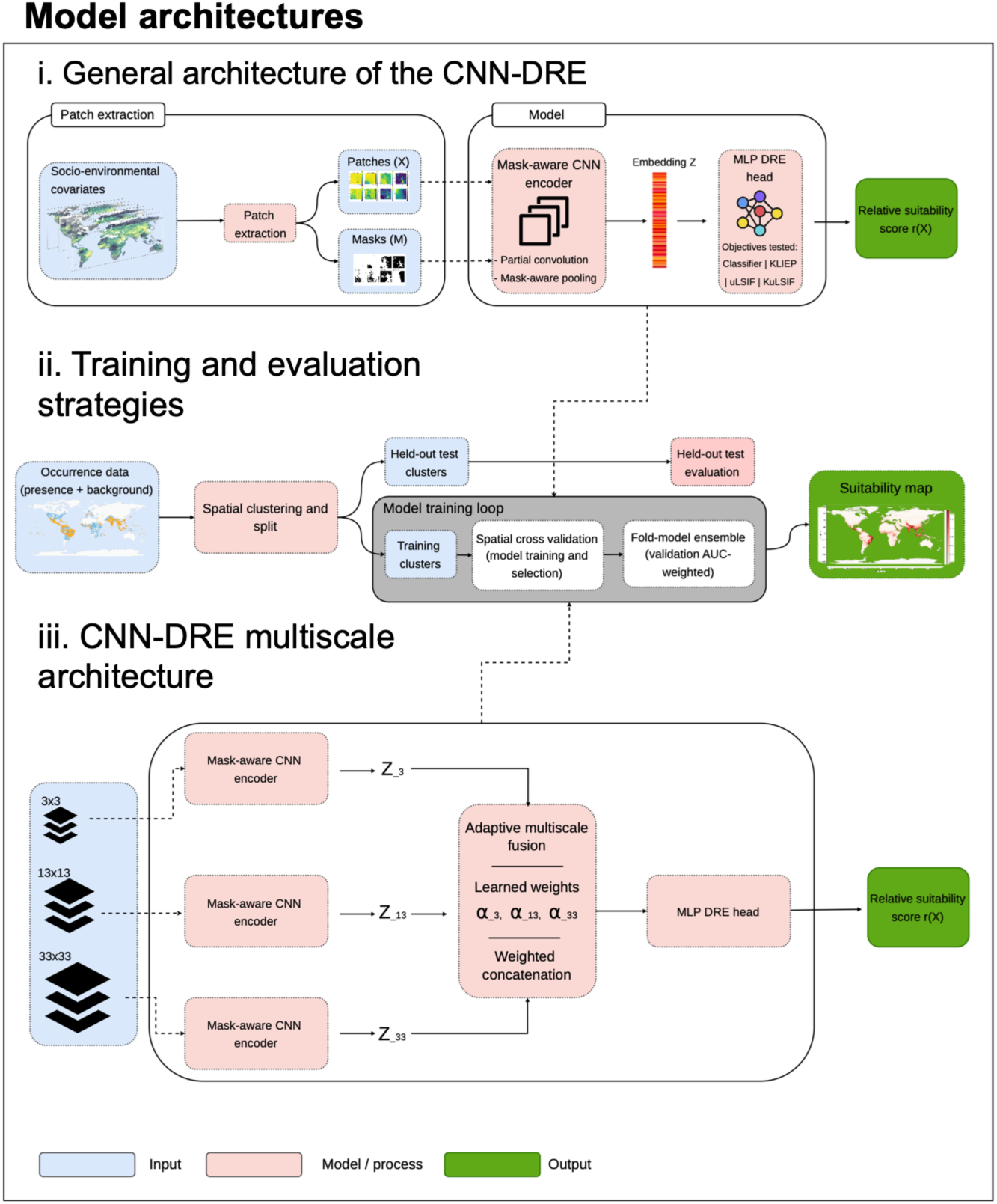

**Supplementary Figure S4.**
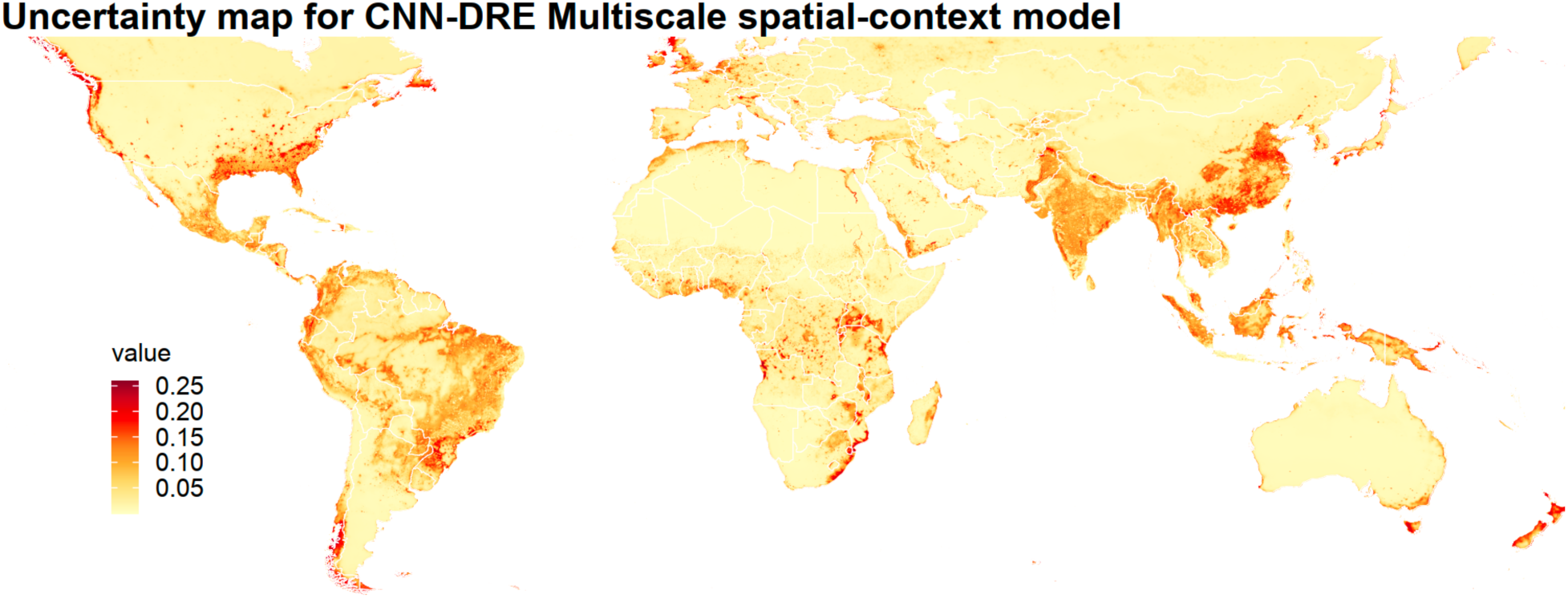
**Multiscale CNN-DRE uncertainty map**

**Supplementary Figure S5.**
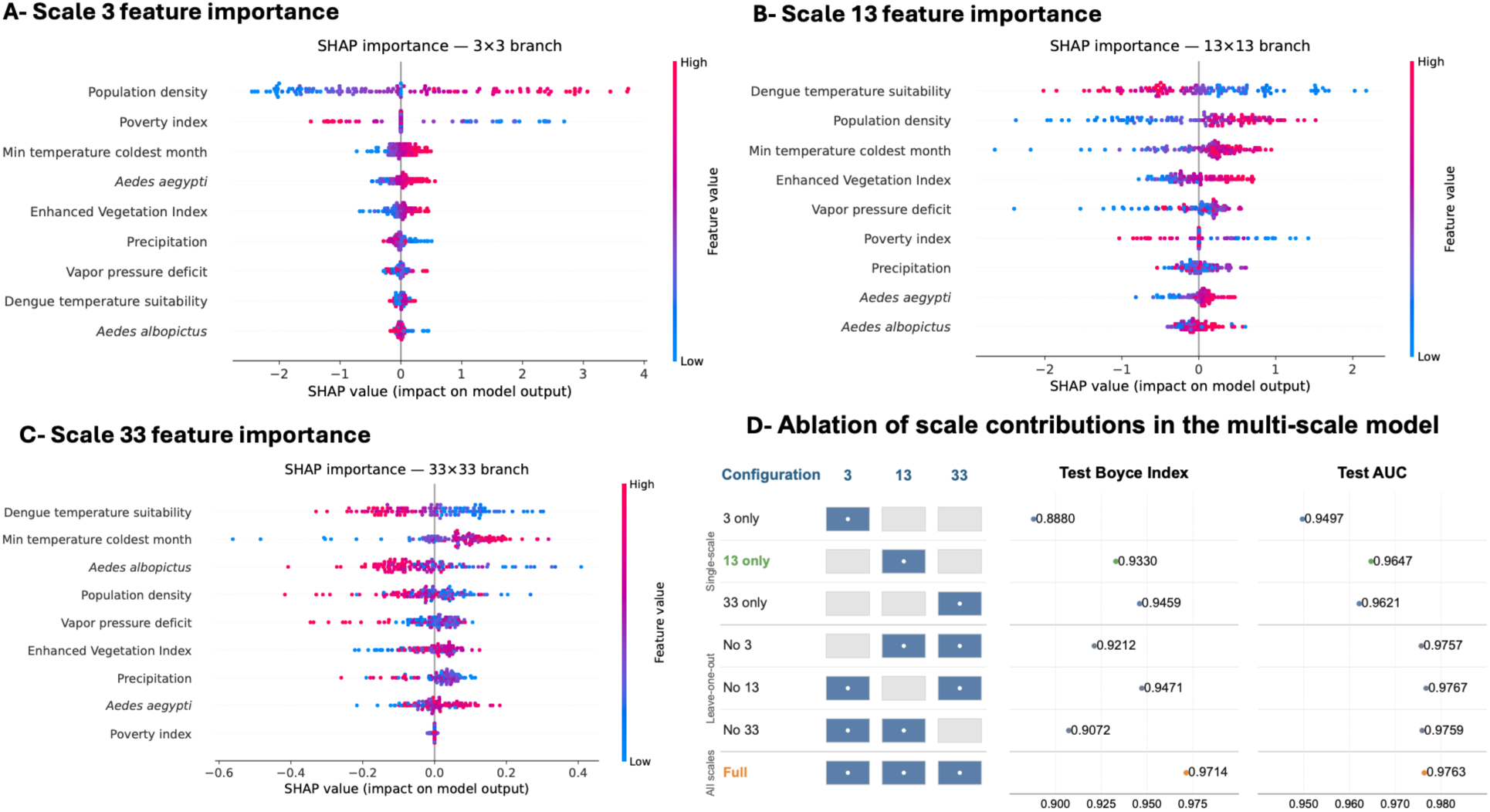
Scale-specific feature attributions and contribution of each branch in the multiscale CNN-DRE model. (A-C) SHAP feature-importance summaries for the 3×3, 13×13, and 33×33 CNN-DRE branches, showing how covariate contributions vary across local, neighbourhood, and broader spatial scales. (D) Ablation of scale contributions in the multiscale model, comparing single-scale, leave-one-scale-out, and full multiscale configurations using held-out test Boyce index and AUC

**Supplementary Figure S6.**
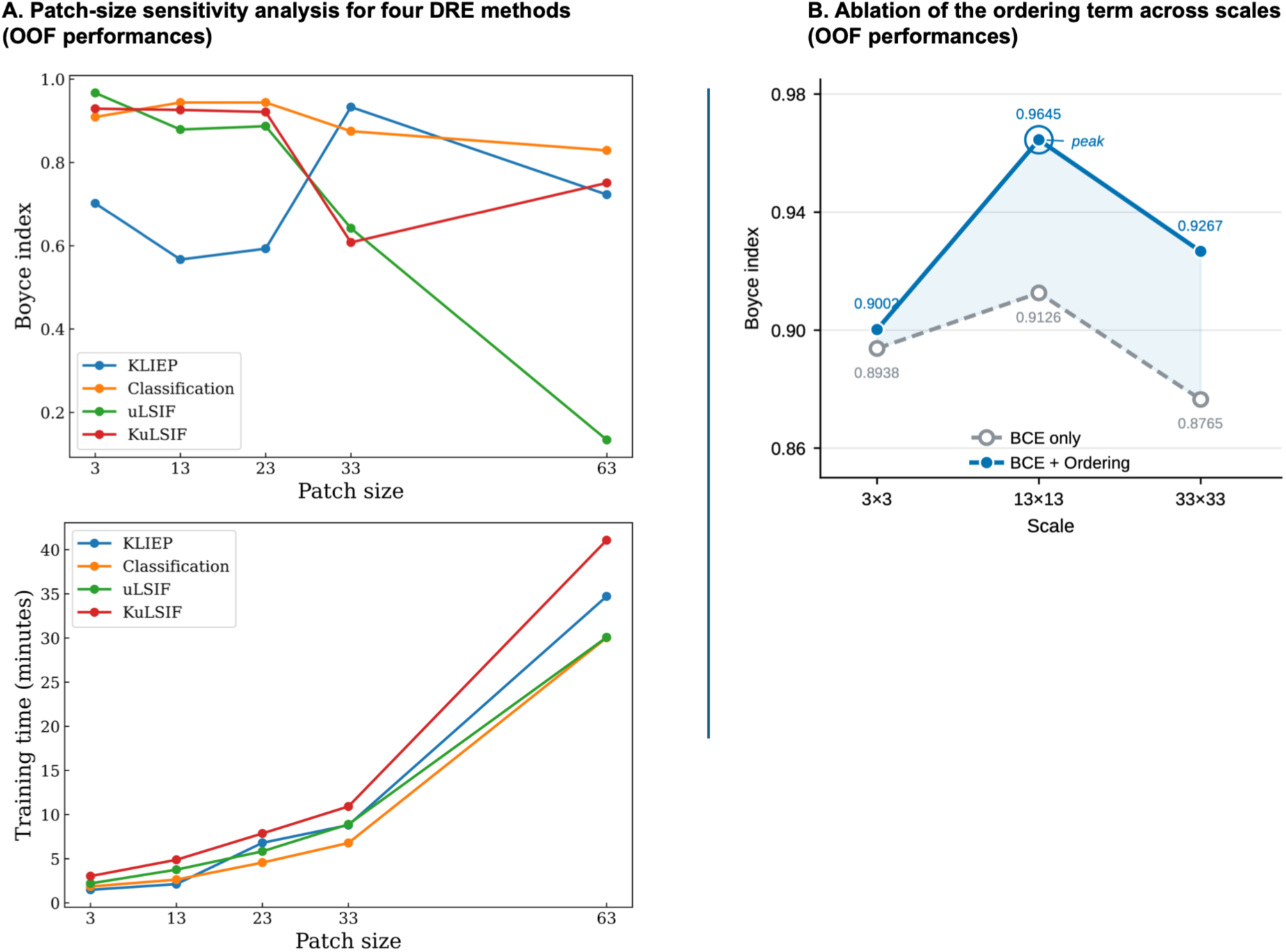

**Supplementary Figure S7.**
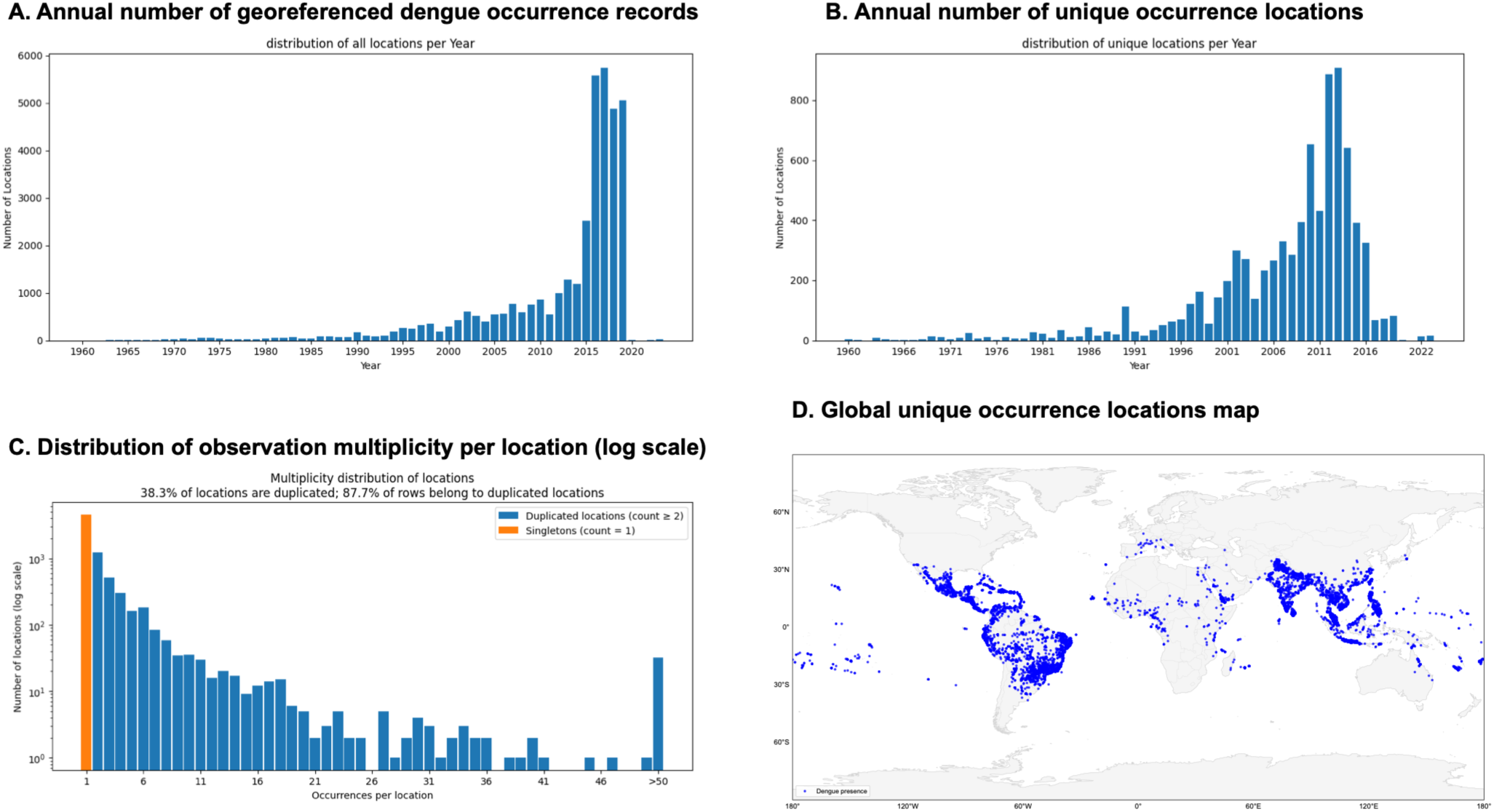
Temporal and spatial georeferenced dengue occurrence records (1960-2023). (A) Annual number of georeferenced dengue occurrence records. Reporting intensity increased markedly after 2000, with most records concentrated in recent years. (B) Annual number of unique occurrence locations after spatial deduplication Although unique sites increased over time, temporal coverage remains uneven, with relatively sparse observations prior to the 1990s and a pronounced peak in the 2010s. (C) Distribution of observation multiplicity per location on a log scale, showing repeated records at some locations and many singleton observations. While 61.7% of locations were reported only once, 38.3% were recorded multiple times; moreover, 87.7% of all records originated from duplicated locations, indicating strong spatial reporting heterogeneity and concentration of observations in a limited number of sites. (D) Global map of unique dengue occurrence locations used for model development and evaluation.

**Supplementary Figure S8.**
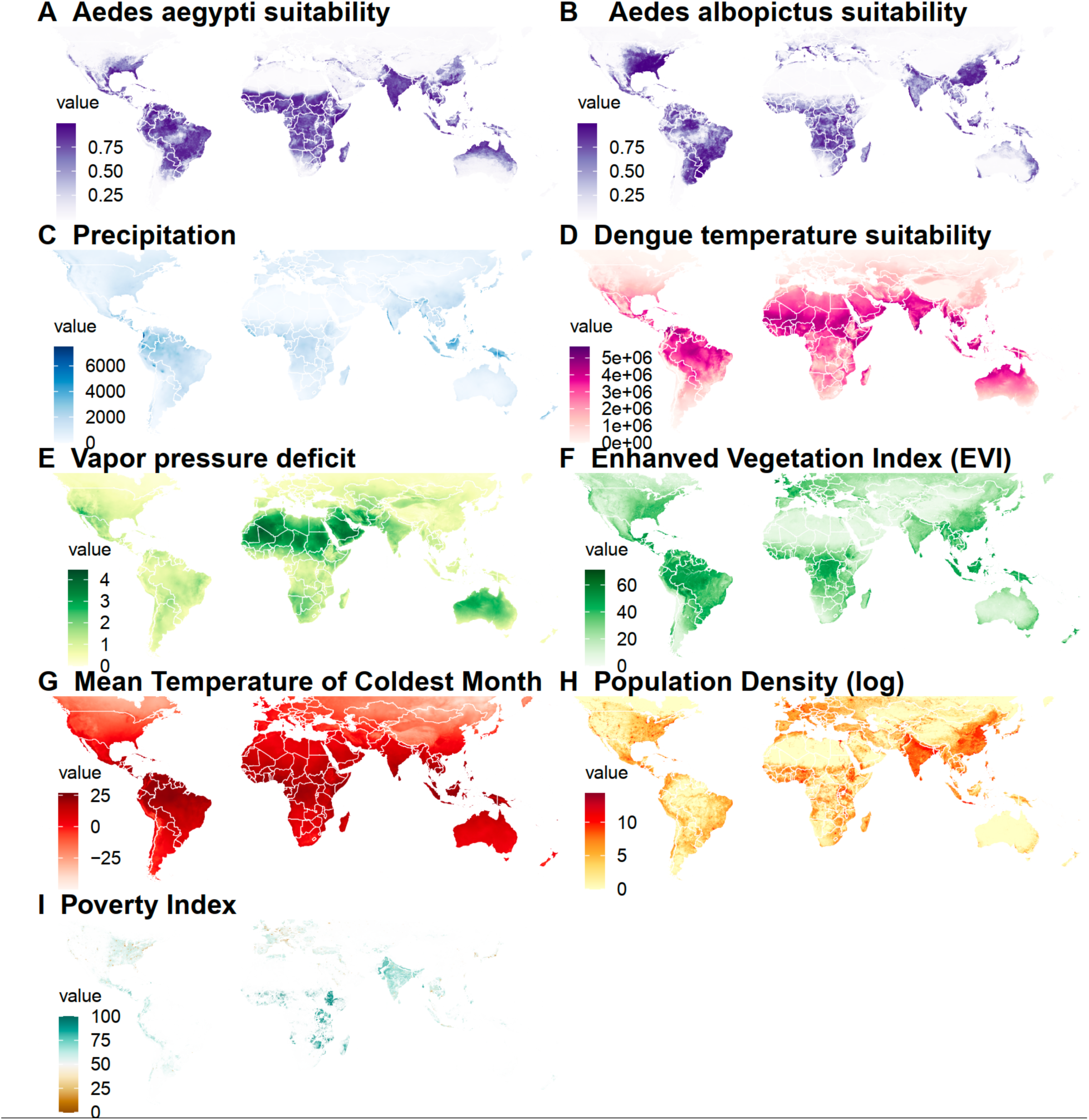
**Global covariate layers used for CNN-DRE dengue suitability modelling**

**Supplementary Figure S9.**
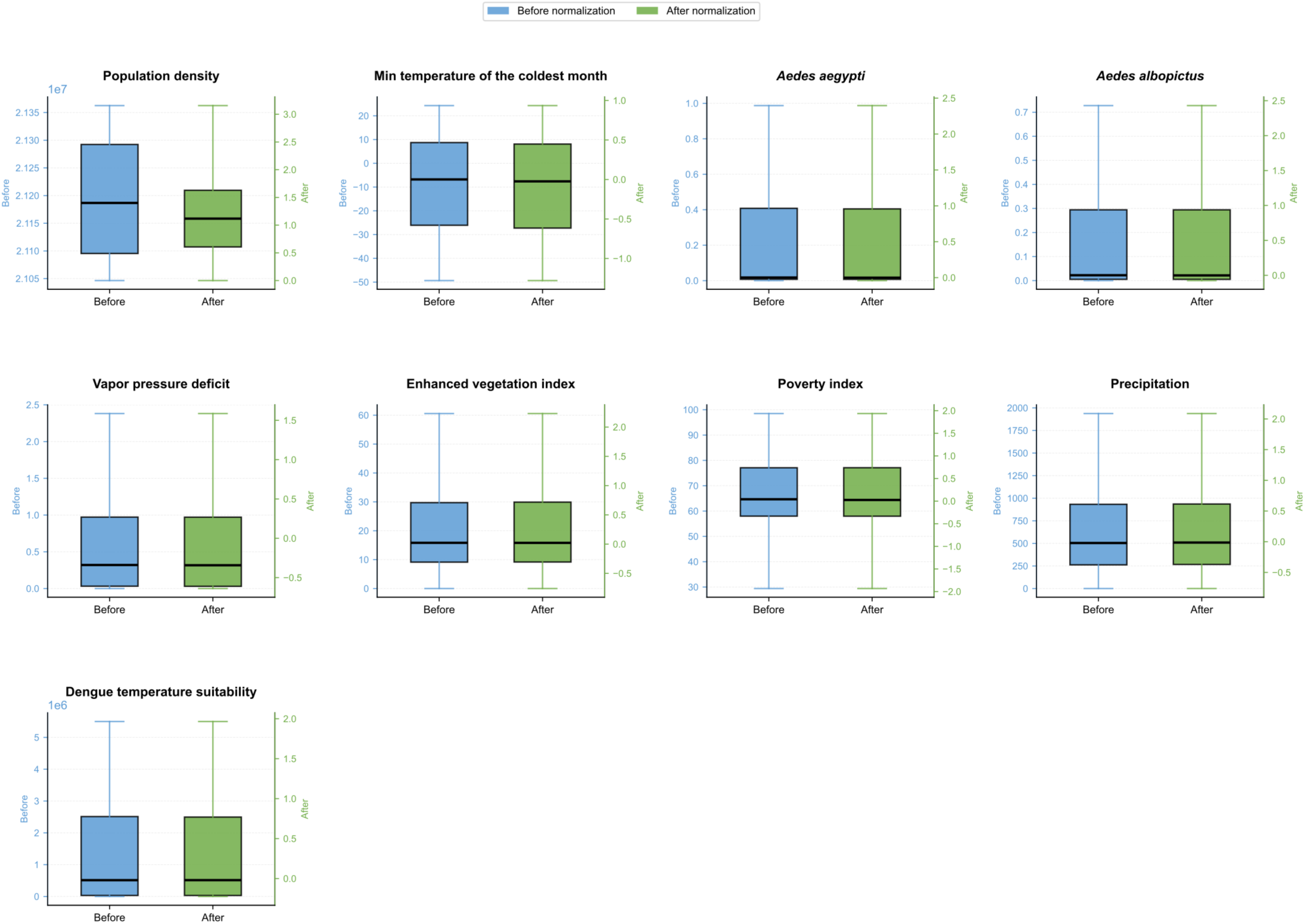
**Covariate distributions before and after normalization**

**Supplementary Figure S10.**
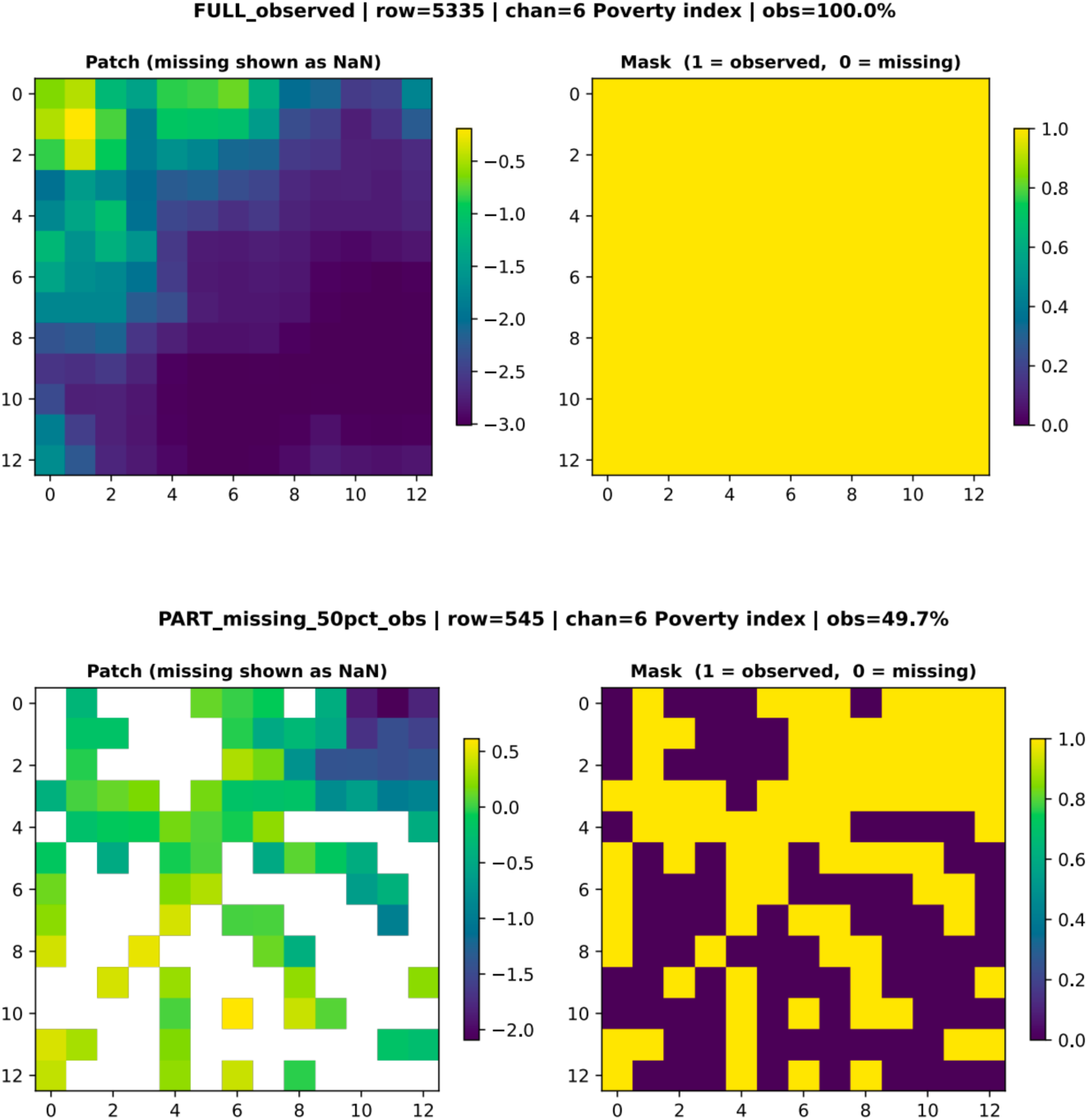
**Example of a patch with its associated mask**

